# Predictive Modelling of Normative Lower Limb Sagittal Kinematics in Young Ghanaian Adults

**DOI:** 10.1101/2025.04.03.25325168

**Authors:** Philip Hotor, Jerry Amegatse, Gabriel Ashitey, Sahene Arkoh

## Abstract

Clinical Gait Analysis (CGA) is a pivotal technique for evaluating pathological conditions, particularly musculoskeletal disorders. However, its efficacy is often hindered by the fact that normative gait data is almost always used worldwide as a basis for CGA, regardless of differences in critical parameters such as BMI, age, gender, and walking speeds. To address this, we developed multiple regression models for predicting lower limb sagittal kinematic waveforms. We recorded anthropometric, demographic, spatiotemporal, and kinematic data from 30 healthy individuals. Leveraging the gait cycle time and joint angles as dependent variables, and BMI, age, gender, and walking speeds as predictors, we developed 46 regression equations. We employed PCHIP utilizing 80% of the kinematic data to reconstruct the waveforms and validated via leave-one-out cross validation. Our models successfully reconstructed hip, knee, and ankle kinematic waveforms, achieving R^2^ ≥ 0.9 and RMSE ≤ 6° from the validation study. P-values < 0.05 as well as the clinical relevance of the predictors were considered during the regression analysis. These outcomes underscore the potential for our approach to be used as the basis to enhance the precision of region-specific gait data predictions, thus facilitating more accurate CGA.

## 1 Introduction

Understanding the biomechanics of human gait is important in various fields, including sports science, rehabilitation, and orthopedics. The gait cycle, comprising various phases such as stance and swing, reflects the complex interplay between muscles, joints, and bones during locomotion (Lu & Chang, 2012). Analyzing gait cycle data from normative databases provides valuable insights into movement patterns, functional impairments, and pathological conditions affecting the lower limbs. Accurate gait analysis enables healthcare professionals to design tailored interventions and monitor the progress of patients with gait-related issues. Sagittal plane kinematics, involving motion along the anterior-posterior axis, are particularly crucial in Clinical Gait Analysis (CGA). Parameters such as hip, knee, and ankle angles during the gait cycle offer information about joint function, stability, and efficiency (Pietraszewski, et al., 2012). Abnormalities in sagittal plane kinematics can indicate musculoskeletal disorders, gait asymmetries, or biomechanical inefficiencies, highlighting the importance of accurate measurement and analysis techniques. In addition, information like the demography, anthropometry, or spatiotemporal from normative gait data (Abdul Aziz Hulleck et al., 2022; Galea et al., 2019) are intrinsic to an individual’s physiology and can significantly affect gait patterns, making their inclusion imperative for a more comprehensive and objective assessment (Al-kharaz & Chong, 2022).

Age is a fundamental factor influencing gait patterns, with distinct age-related changes in gait well-documented. These changes underscore the need for age-specific normative databases, which are not always readily available (Vervoort, 2020; Rowe et al., 2021). Gender is another factor contributing to variations in gait patterns, with females typically displaying distinct gait characteristics compared to males. These gender-specific differences extend to kinematic features of gait (Rowe et al., 2021). Walking speed, another fundamental gait parameter, affects various biomechanical variables, including joint kinematics. Changes in walking speed can lead to significant deviations in gait parameters, making it a critical consideration in gait analysis (Sun et al., 2018; Florent Moissenet et al., 2019). Furthermore, Body Mass Index (BMI), a measure of an individual’s body mass relative to their height, is intricately connected to gait parameters. Variations in BMI can result in distinct gait patterns (Shadi Eltanani et al., 2023).

Regression analysis is a powerful technique for predicting biomechanical metrics based on various predictors such as anthropometric measurements, demographic factors, and physiological parameters. By fitting regression models to experimental data, researchers can elucidate the relationships between predictors and response variables, enabling predictive modeling and hypothesis testing. Robust regression techniques enhance the reliability of predictions by accounting for outliers and heteroscedasticity in the data. Statistical analysis like the mean and standard deviation metrics of gait cycle data allows for quantification of variability, consistency, and trends in joint angles, velocities, and accelerations. These analysis reveals relationships between joint angles and gait cycle phases. Such analyses also aid in identifying normal and abnormal gait patterns, informing clinical assessments and rehabilitation interventions (Mikos et al., 2018). Despite advancements in gait analysis techniques, several challenges persist, including data variability, model complexity, and limited generalizability. Future research directions may involve exploring nonlinear regression models, incorporating machine learning algorithms, and integrating multi-modal data sources for comprehensive gait analysis. Addressing these challenges will enhance the accuracy, reliability, and clinical utility of gait analysis methodologies, ultimately improving patient care and biomechanical research outcomes.

While previous research has explored multilinear regression models to predict normal gait patterns, many of these models were developed using data from the Western population (Florent Moissenet et al., 2019). However, the gait system is inherently unique to each individual, making such models prone to errors when applied to new dataset (Strongman & Morrison, 2020; Eni Halilaj et al., 2018). To address this limitation in the African context, our study seeks to predict and analyze the lower limb sagittal kinematic waveforms based on the BMI, age, gender, and walking speed of the young Ghanaian adult.

In the subsequent sections, we will delve into the methods employed for data collection, analysis, and modeling. We will present the demographic and anthropometric data of our participants, as well as spatiotemporal gait parameters. The kinematic data and key points used in our models will be thoroughly discussed. Furthermore, the results of our study, including the validation of our predictive models, will be presented. Our research represents a significant step forward in the field of clinical gait analysis, offering a comprehensive and more accurate method for evaluating gait patterns while considering individual variations. By integrating these gait predictors, we aim to enhance the precision and reliability of gait assessments, ultimately contributing to better CGA.

## 2 Methods

### 2.1 Participants

This study involved a carefully selected group of 30 healthy young adults (16 female, 14 male) between 17 and 25 years, all students from the School of Engineering Sciences at the University of Ghana. The inclusion criteria required participants to have no prior gait disorders, ensuring that the sample was homogeneous regarding gait mechanics. The data recording process was briefly explained to the participants and they were made to fill a consent form which was approved by the University of Ghana local Ethics Committee of Basic and Applied Sciences prior to their inclusion. The protocol was conformed to the Declaration of Helsinki for human experiments (World Medical Association, 1974).

### 2.2 Experimental Protocol

Demographic (age, gender), anthropometric (height, weight), sagittal kinematic waveform (hip, knee, ankle) and spatiotemporal (walking speed, stance/swing phase, double step length, range of motion) data were collected. Table 2 and 3 as well as Figure 1 show details of the spatiotemporal data. The height and weight of the participants were measured using a stadiometer and a weighing scale (CMS Weighing Equipment Ltd., London NW1, England), respectively. To measure the height each participant was asked to remove any footwear and stand against a flat surface such as a wall with no protrusion. We ensured that the feet of the participants were flatly placed on the floor and together, legs were straight, arms by the side, and shoulder leveled. To measure the weight, each participant was asked to remove any footwear or heavy clothing and stand in the center of a scale placed on a flat hard surface.

**Table 1:**
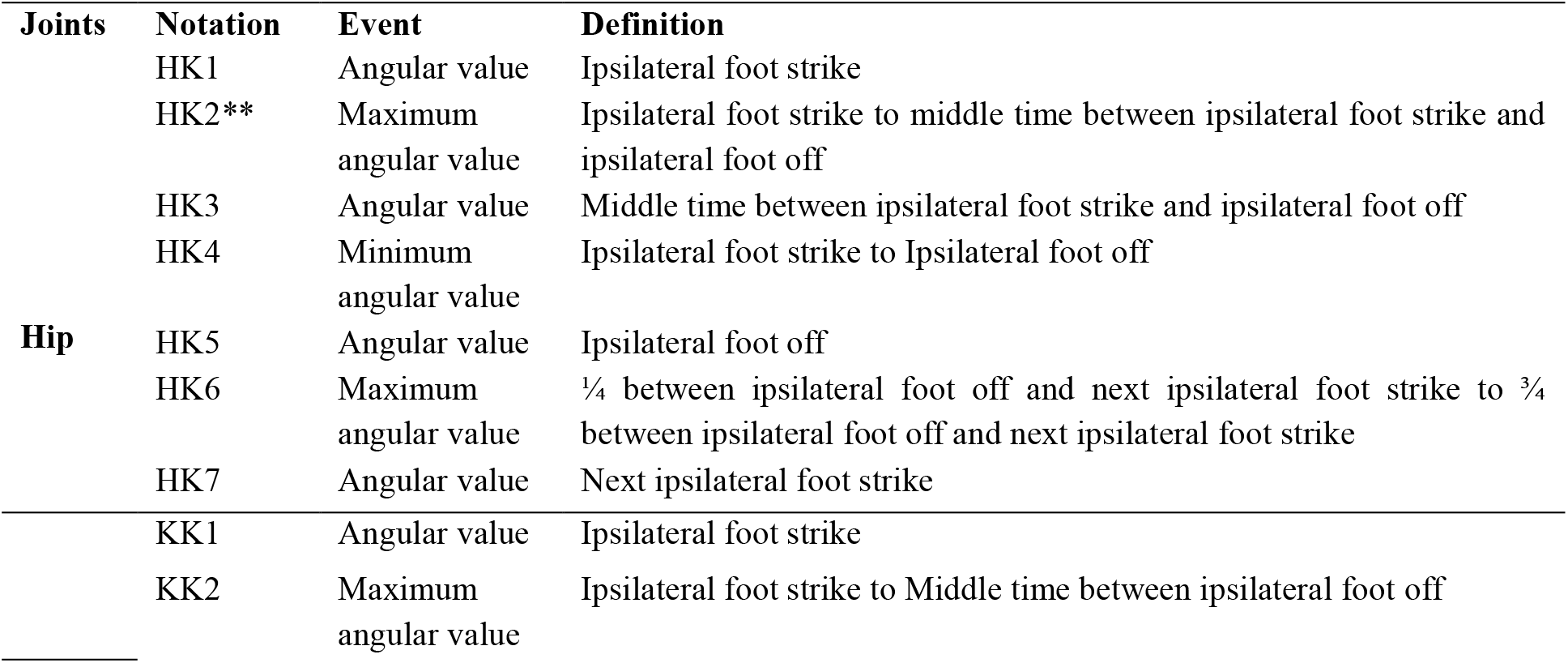
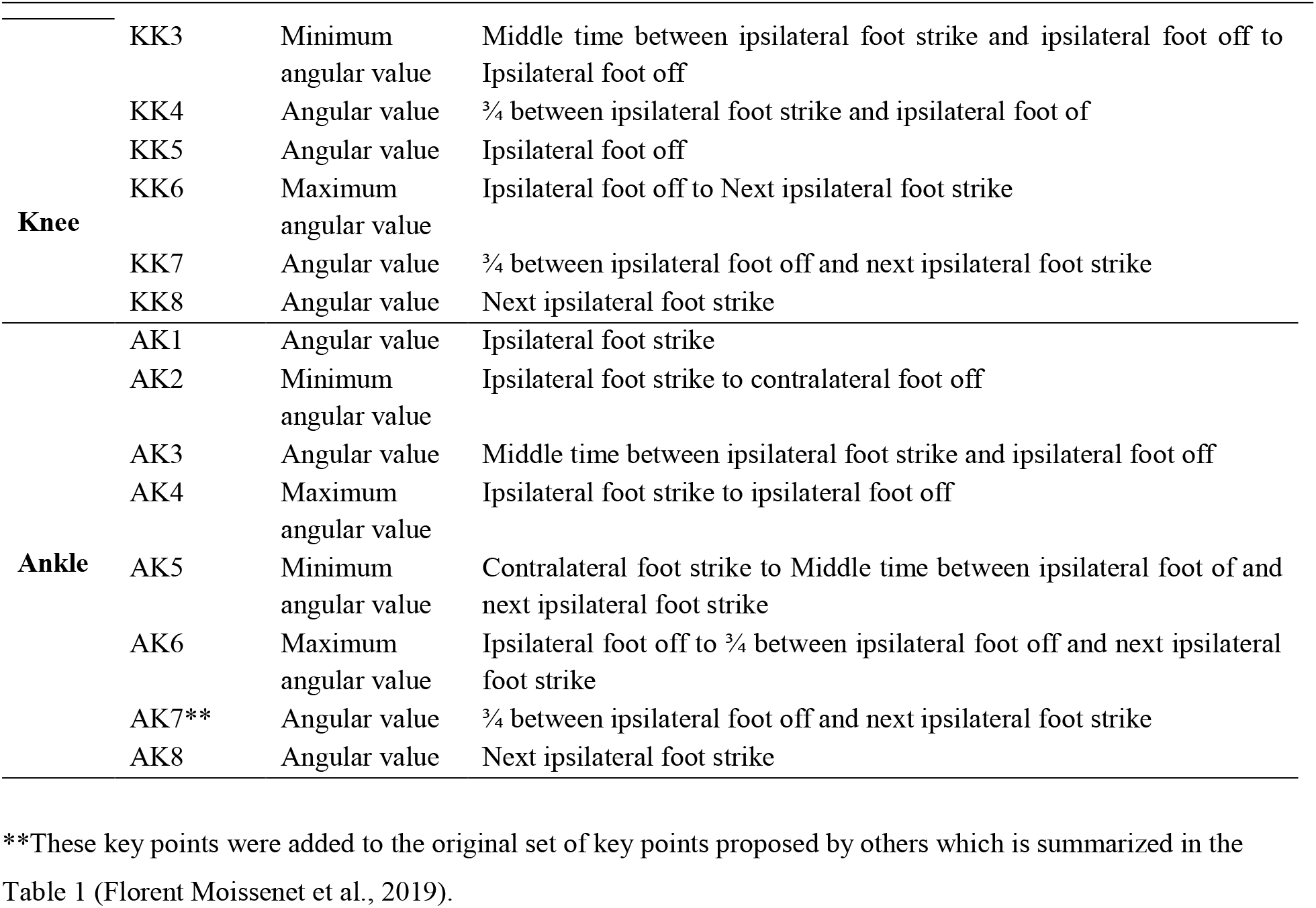
Key points extracted for hip, knee, and ankle joints during movement analysis.

**Table 2:** The distribution of demographic data, including age and gender, among the 14 males and 16 females. The anthropometric data, including mean weight, height, and BMI, were also recorded, along with the standard deviations (SDs) and coefficient of variation.

| Parameter | Male | Female | Mean(SD) | CV (%) |
| --- | --- | --- | --- | --- |
| Age (yrs.) | 20.93 (2.34) | 20.63 (1.75) | 20.77 (2.01) | 9.81 |
| Weight (kg) | 67.19 (16.86) | 63.74 (12.39) | 65.35 (14.49) | 22.17 |
| Height (m) | 1.74 (0.09) | 1.60 (0.09) | 1.66 (0.12) | 6.94 |
| BMI (kg/m<sup>2</sup>) | 22.06 (3.65) | 25.14 (5.28) | 23.7 (4.82) | 20.34 |

**Table 3:**
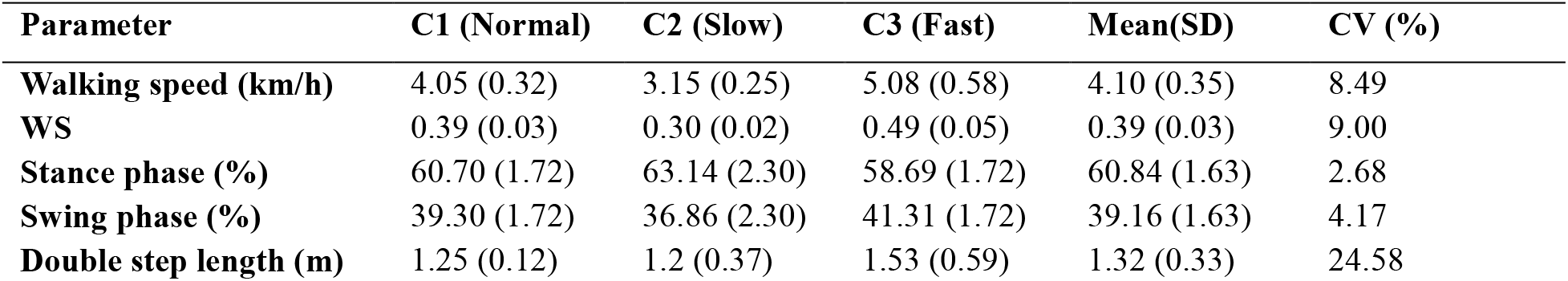
The mean, standard deviations, and coefficient of variability values of spatiotemporal gait parameters of participants under different walking conditions.

**Figure 1:**
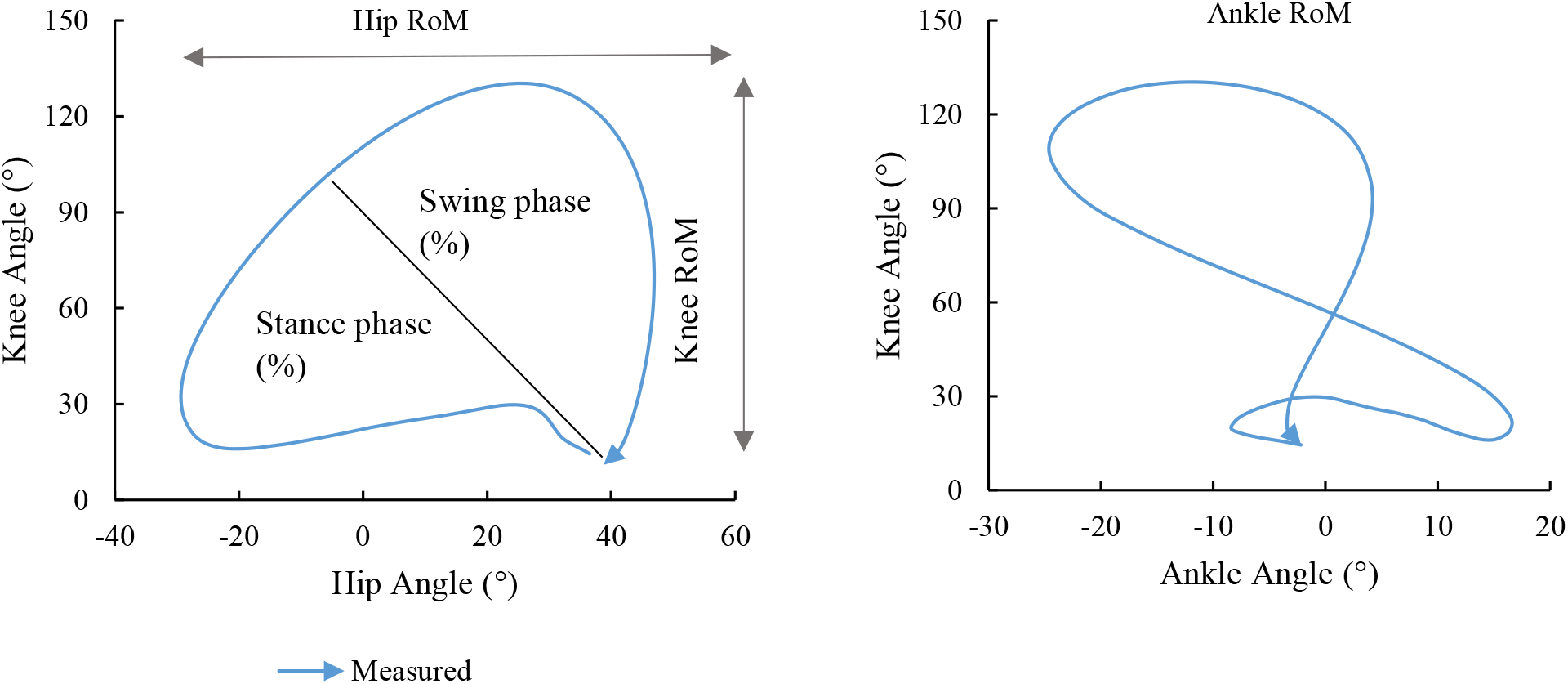
The cyclograms estimate and illustrate the range of motion in the hip flexion/extension, knee flexion/extension, and ankle plantar/dorsi flexion as well as the stance and swing phase of the gait cycle.

An instrumented gait analysis set up which comprised of a treadmill (Domyos W500 treadmill, Decathlon, China), two high speed cameras (Blue Fox3 and CougarX), and a motion capture software (Simi Aktisys v 2.1.4.0; Simi Reality Motion Systems GmbH, Unterschleissheim, Germany) was used to capture the sagittal kinematic waveforms for the hip, knee, and ankle joint angles as well as double step length, duration of the stand and swing phases, and range of motion of each participant at 100Hz. The motion of the participants were captured within 2/3 of the camera frame. Each participant underwent a familiarization period of at least three minutes on the treadmill before the actual data acquisition. Using anatomical palpation, five active colored LED markers were stuck on the participant’s joints in accordance with the recommendation by Celine Schreiber (Schreiber & Florent Moissenet, 2019). The markers were placed on the acromion of the shoulder joint, the trochanter major of the hip, and the lateral femoral epicondyle of the knee. The remaining markers were placed on the lateral malleolus of the ankle and the fifth metatarsal basic joint of the forefoot. The participants were asked to walk on the treadmill so that their motion were recorded for three trials with a minute break between each trial. The data was recorded at self-selected normal (*C*1), below the normal (*C*2), and above the normal (*C*3) walking speeds.

### 2.3 Preprocessing

The motion capture data consist of walking speed, double step length, gait cycle time (stance phase, swing phase), range of motion, hip, knee, and ankle joint angles. To obtain the kinematic data, a single analysts manually specified 8 key events of the gait cycle (initial contact, loading response, mid-stance, terminal stance, pre-swing, initial swing, mid-swing, terminal swing) and corrected marker dislocations using the Simi motion capture software system. The walking speeds were converted into dimensionless walking speed, *WS* by dividing the raw walking speed (*m. s*^−1^) by the square root of the product of the leg length (*m*) and the gravitational constant (*m. s*^−2^) (Hof, 1996). The leg length, distance from the hip joint to the ground, was computed by multiplying 0.53 by the total body height (*m*) (Karimi & Jahani, 2012).

Kinematic waveforms of hip, knee, and ankle were time-normalized (0 − 100% with increments of 1%) via interpolation using a cubic spline function in MATLAB R2021a (Mathworks Inc., Natick, MA, USA). Key points for the hip, knee, and ankle gait features (gait cycle time, angle) were extracted automatically using custom-written MATLAB script and denoted as *HKi, KKi, AKi* (*i* can be 1, 2, 3, 4, 5, 6, 7, *or* 8) respectively based on the definitions in Table 1. According to the definitions, we used 7 key events for hip and 8 for both knee and ankle gait features (Florent Moissenet et al., 2019). Although using more key points would have resulted in reconstructed waveforms that were considerably closer to the originals, key points with biomechanical and clinical significance were chosen instead, as proposed by others (Galea et al., 2019).

### 2.4 Regression Analysis

Multiple linear regression analysis were performed on our dataset which contains multiple predictors and response variables. The predictors include WS, BMI, gender, and age at their mean values. Females were coded as 0 and males as 1 such that positive regression coefficients means positive effect for males and negative effects for females. While the response variables are gait cycle(%) and angle(°). For each response variable, three types of regression models were fitted, namely

1. Linear Model (LM), which is a basic linear regression model
2. Stepwise Linear Model (SLM), also known as a stepwise regression model that iteratively adds or removes predictors based on their statistical significance
3. Robust Linear Model (RLM), a regression model using robust fitting techniques to handle outliers and heteroscedasticity.

To obtain the regression coefficients, we defined the robust linear model in the form of Equation 1. Equation one applies the RLM to the predictor and response vector and returns the coeffecients and RLM statistics, reported in Table 4. In Equation 2, the variables *Ki* are the new predicted response variables, *β*_0_ is the intercept and *β*_1_, *β*_2_, *β*_3_, *β*_4_ are the coefficients for the predictors. The predicted values, *Ki* for each response variable were calculated using the coefficients from the robust linear model and the predictors.

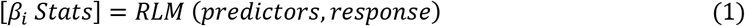

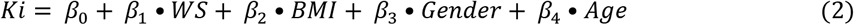

**Table 4:**
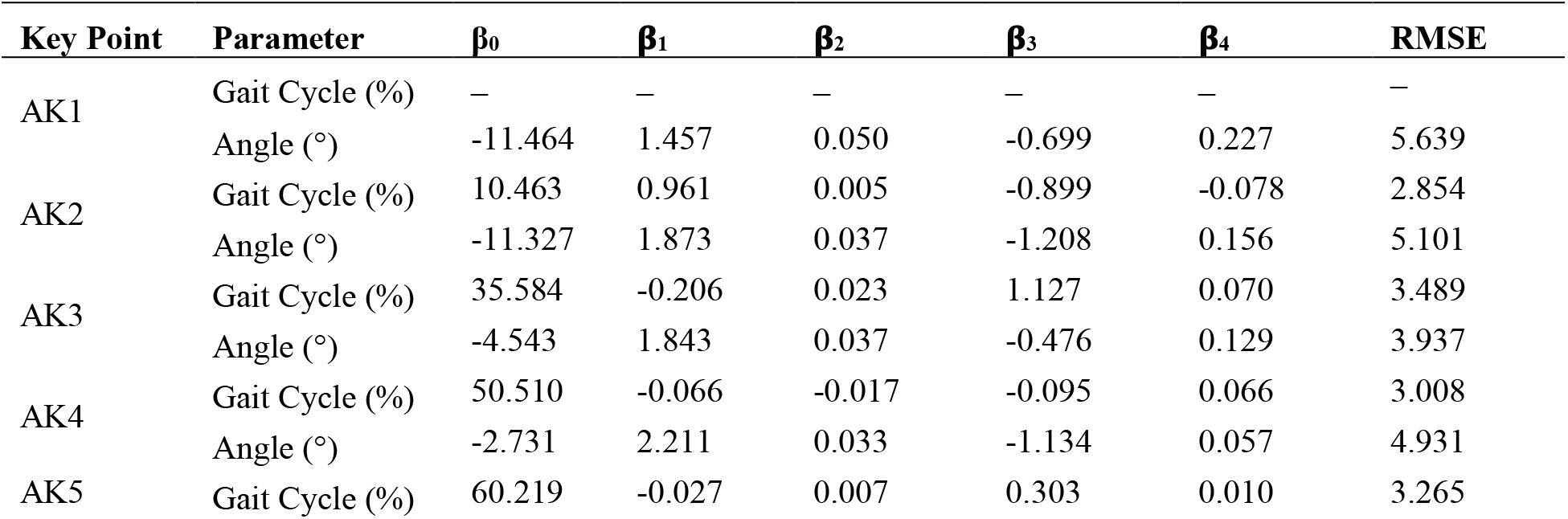
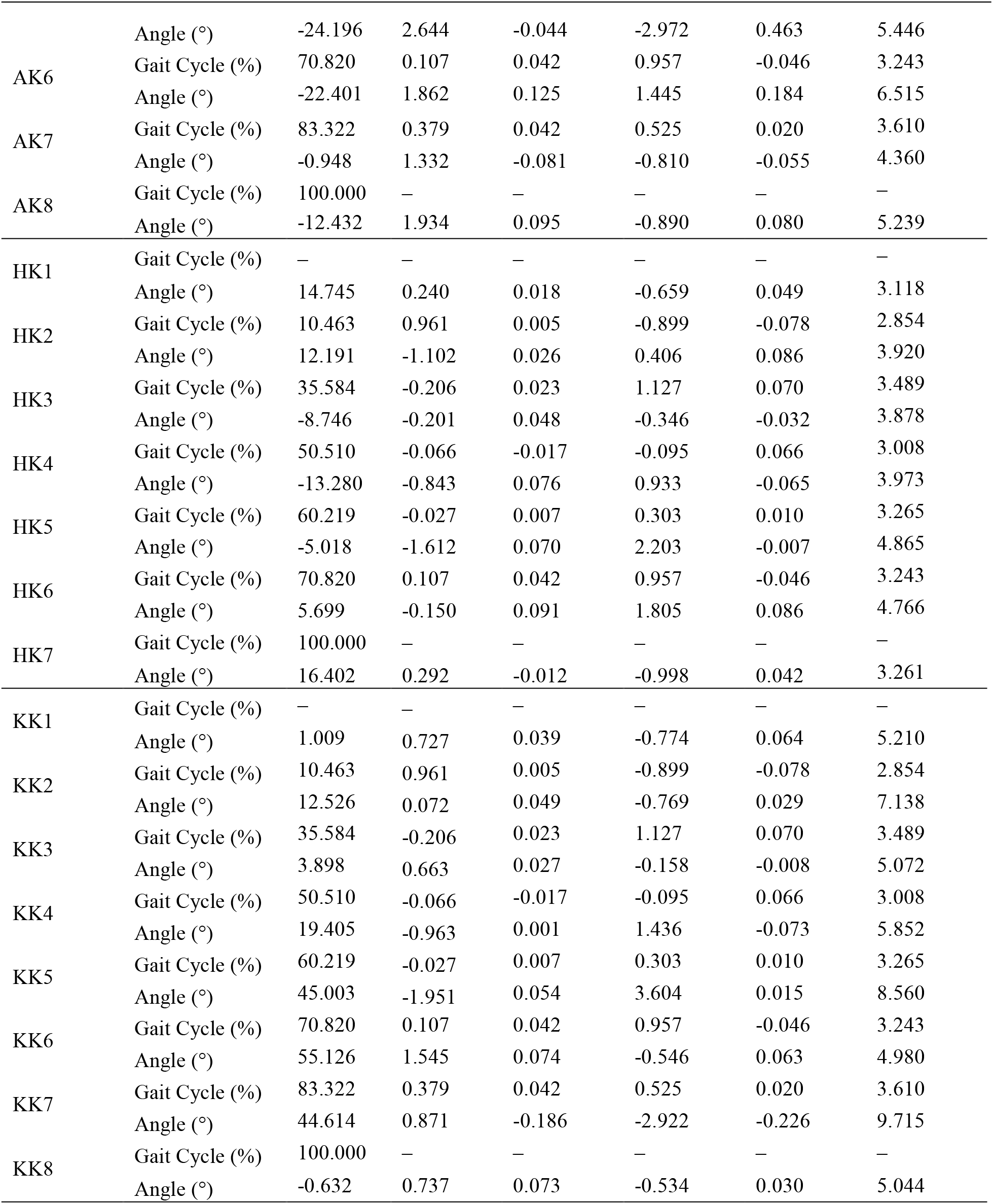
Regressors for the hip, knee, and ankle waveforms.

### 2.5 Waveform Reconstruction

The predicted values, *Ki* were used to reconstruct the waveforms for hip, knee, and ankle. To ensure smooth transitions and continuity in the data, the first and last value of each data points for hip, knee, and ankle data fields are set to be equal. A custom MATLAB script using Piecewise Cubic Hermite Interpolating Polynomial (PCHIP) algorithm was developed to reconstruct and optimize the waveforms. PCHIP constructs cubic polynomial between each pair of key point with specified derivatives (slopes) at the key points. This generates a shape-preserving smooth waveform between key points (Fritsch & Carlson, 1980; Kahaner et al., 1988). Equations 3 to 6 explains the PCHIP algorithm.

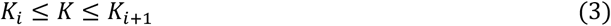

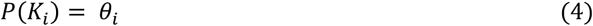

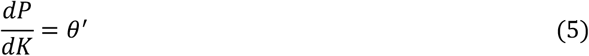

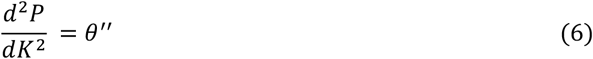

Equation 1 represent a pair of key points where *P*(*K*) is a cubic Hermite interpolating polynomial. In Equation 3, *P*(*K*) interpolates angle, *θ* so that the slopes, *θ*^*′*^ and *θ*^*′′*^ are continuous over the gait cycle. The slopes at the *K*_*i*_ are chosen such that *P*(*K*) preserves the shape of the data and continuous with respects to gait cycle, thus respects monotonicity. Therefore, on intervals where the data is monotonic, so is *P*(*K*), and at points where the data has a local extremum, so does *P*(*K*). For instance, at *HK*_4_ (local minimum) and *HK*_6_ (local maximum), the data is monotonic and so does *P*(*K*). Similar trends can be observed in *KK*_*i*_ and *AK*_*i*_ data points. This method is chosen for its ability to preserve the shape and features of the data, avoiding oscillations common with other interpolation methods (KAYA, 2014).

### 2.6 Statistical Analysis

All statistical analysis were conducted using MATLAB R2021a and Excel 2013 software. The reconstructed waveforms were validated using leave-one-out cross validation. The coefficient of determination, R^2^ and the root mean squared errors, RMSEs of the reconstructed waveforms were computed. The waveforms were also tested via leave-one-out cross validation technique (Florent Moissenet et al., 2019). The reconstructed waveform was superimposed against the three conditions of WS (C1, C2, C3) in our database. The effects of WS on the gait kinematic waveforms for the knee, hip, and ankle joints were analyzed by computing R^2^ and RMSE values. We were able to discuss the R^2^ and RMSE values in details in results and discussion sections. The p-values < 0.05 as well as the clinical relevance of predictors were considered for both SLM and RLM analysis.

## 3 Results

### 3.1 Demographic and Anthropometric Data

The distribution in Table 2 shows demographic data, including the age and gender of participants (sample, *n is* 30) in this study. The anthropometric data (weight, height, and BMI) were also recorded and the coefficients of variation (CV) was computed. The CV indicates the relative variability of each parameter within the sample.

### 3.2 Spatiotemporal Data

The spatiotemporal data in Table 3 shows parameters recorded during motion capture. Parameters include average walking speeds, stance phase, swing phase, and double step length of participants over three different walking conditions (*C*1, *C*2, *C*3). The means, SDs, and CVs of the parameters were also recorded.

The cyclograms illustrates the hip-knee and knee-ankle coordination of all participants and were used to access their range of motion (RoM). The cyclograms were constructed in a clockwise direction for a single gait cycle. The gait cycle is divided into the swing and stance phase with the swing and stance phase contributing to 39.16% (1.63%) and 60.84% (1.63%) respectively for our dataset. The RoM are illustrated as hip, knee, and ankle RoM in Figure 1. The RoM for the hip, knee, and ankle are 76.26° (26.17°), 119.28° (39.66°), and 41.23° (10.84°) respectively.

Below is Figure 3, which shows the time-normalized waveforms of (a) the hip (b) the knee and (c) the ankle for a single gait cycle. The blue curve represents the mean curve for 24 participants sampled randomly from our dataset and the shaded light blue area shows the data points of one standard deviation within the mean of the sampled data. Table 4 summarizes the 46 regression equations including regressors for each predictor of the various key points defined in Table 1. The RMSE values for each key point are also presented.

### 3.3 Kinematic Data

**Figure 1:**
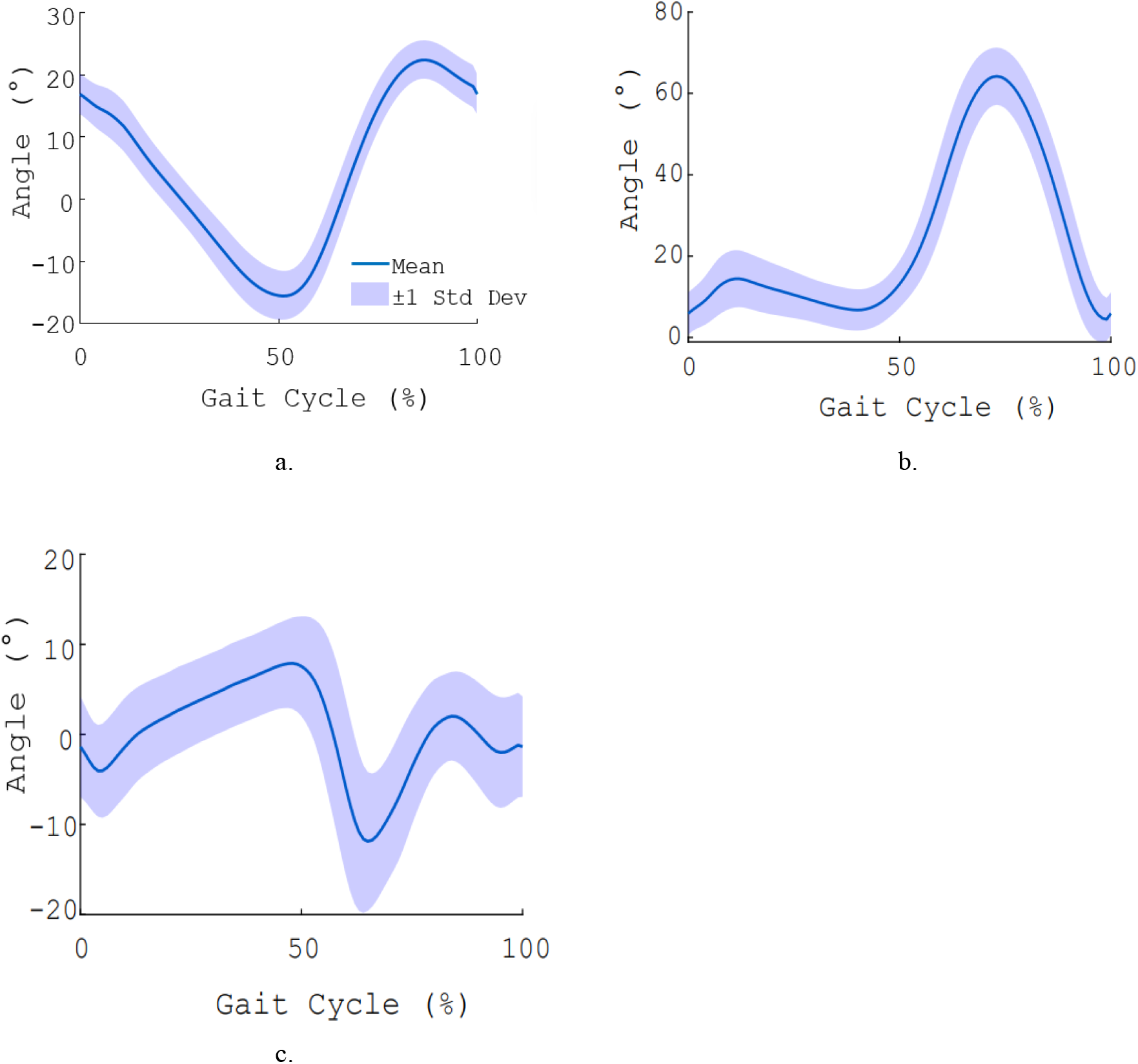
Time-normalized waveforms of (a) hip, (b) knee, and (c) ankle of one gait cycle for 24 participants. The blue curve represent the mean curve and shaded light blue area is the (±1) standard deviation of the mean.

Below is Figure 4, which shows the time-normalized waveforms of the measured (blue curve), predicted (black curve), and key point (red markers) data of (a) hip, (b) knee, and (c) ankle waveforms. The predicted waveforms were obtained with the following parameters: BMI, age, gender and dimensionless walking speed with mean values 23.7 kg/m^2^, 20.8 years, (0 or 1) and 0.39, respectively. R^2^ and RMSE values computed for measured and predicted hip, knee, and ankle waveform 0.981, 0.952, 0.900 and 3.02°, 5.06°, 6.01° respectively.

**Figure 2:**
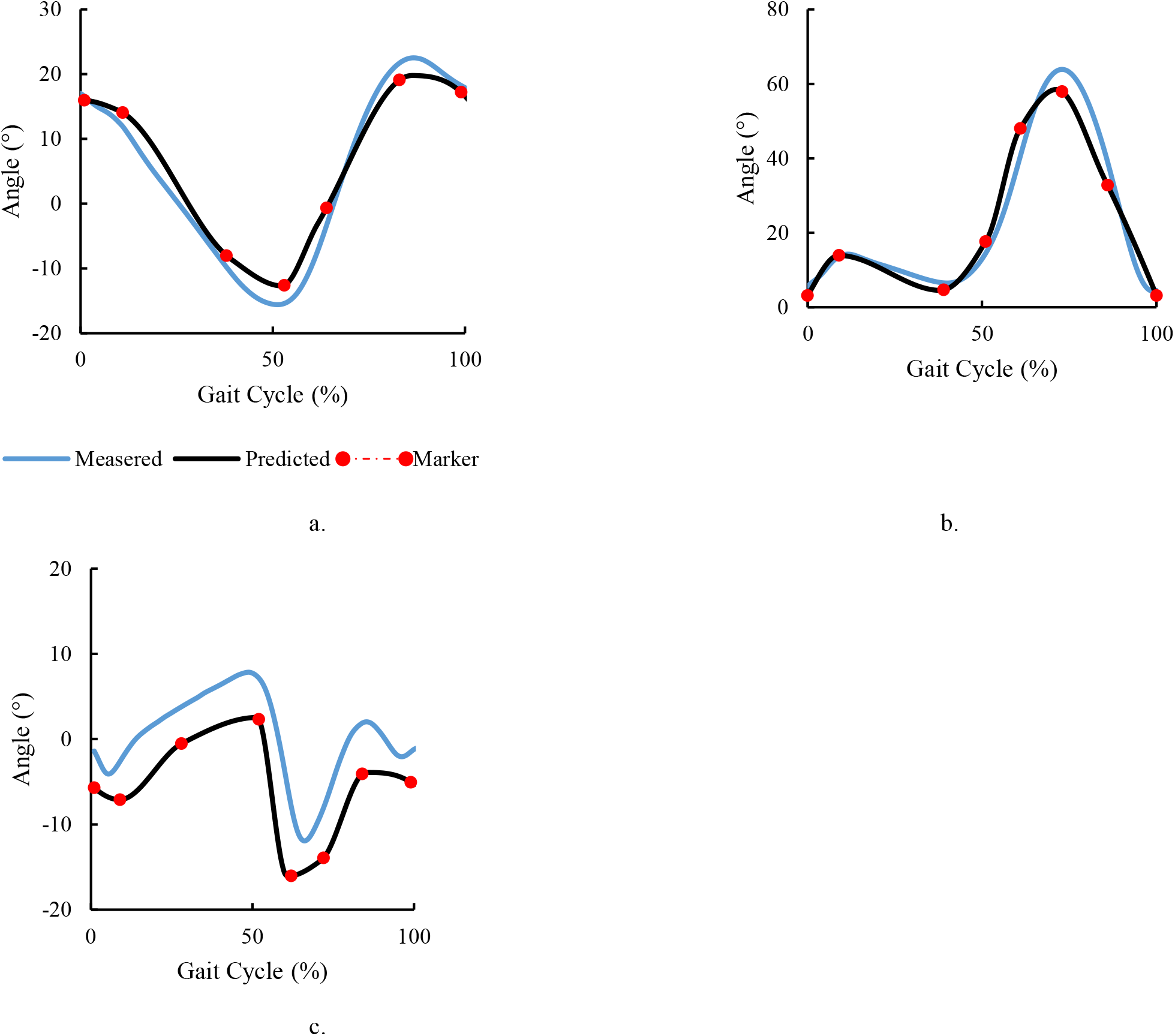
A time-normalized waveforms of the measured (blue curve) for *n* = 6 and predicted (black curve) for *n* = 24 of the (a) hip, (b) knee, and (c) ankle of a participants with the following parameters/predictors: BMI, age, gender, and WS with mean values of 23.7 kg/m^2^, 20.8 years, (0 or 1) and 0.39 respectively. The red markers on the predicted curves are the various data points for the key points.

In Figure 5, The predicted curve was superimposed against curves of C1, C2, and C3 for n = 6. After cross-validation, the R^2^ and RMSE for the three different speed conditions were computed for hip, knee, and ankle respectively. R^2^ for C1 is 0.986, 0.954, 0.913 and RMSE is 3.10°, 5.26°, 3.55°. R^2^ for C2 is 0.962, 0.932, 0.963 and RMSE is 3.04°, 5.10°, 10.07°. R^2^ for C3 is 0.983, 0.699, 0.856 and RMSE is 3.61°, 4.69°, 8.14°.

**Figure 3:**
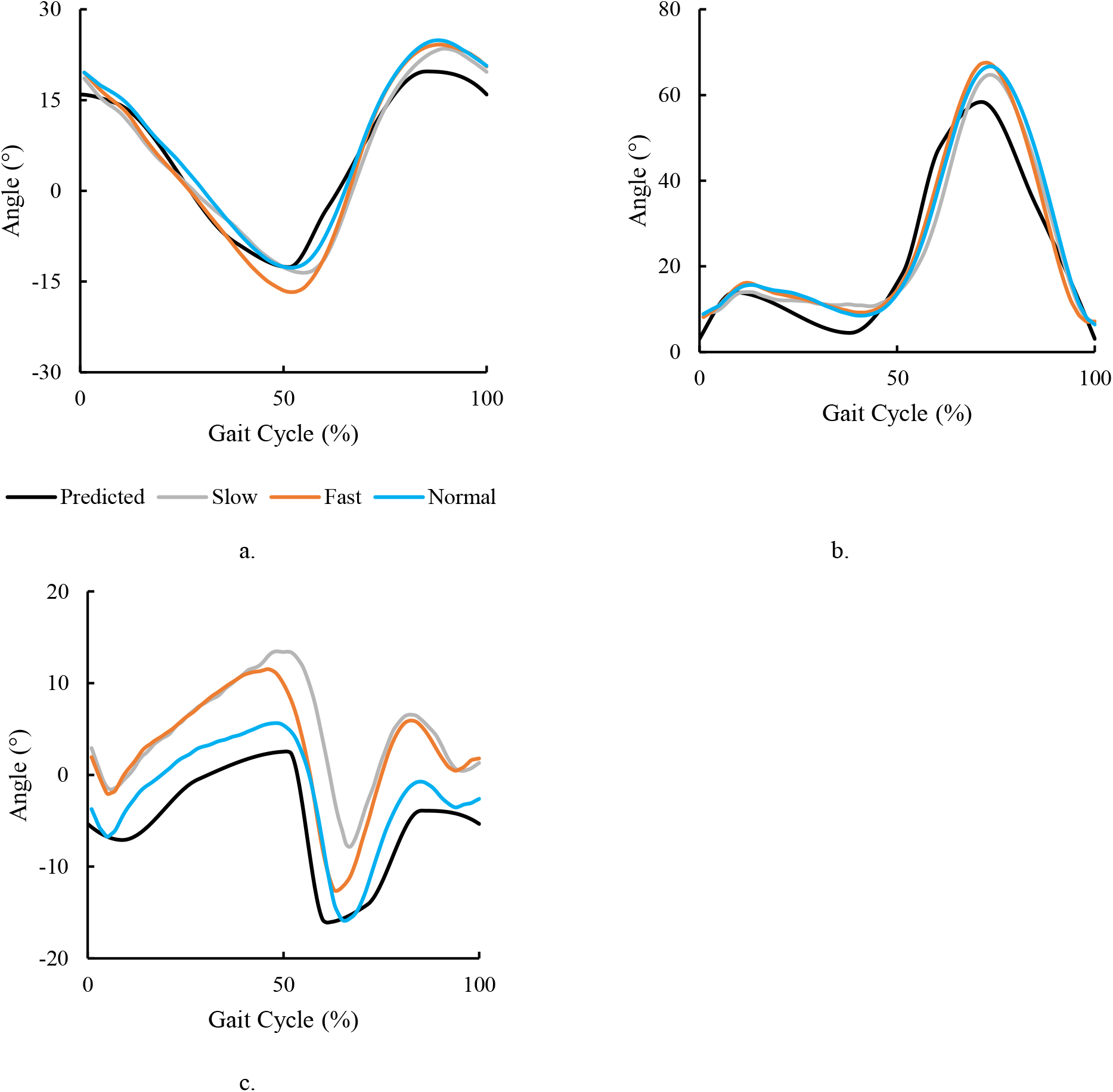
A cross validation test of the time-normalized waveforms of the measured curves and predicted (black curve) of the (a) hip, (b) knee, and (c) ankle of a participants self-selected normal, slow, and fast walking conditions.

## 4 Discussion

Our study aimed to develop predictive models for lower limb sagittal kinematic waveforms in young Ghanaian adults by incorporating key individual characteristics like the BMI, age, gender, and walking speed. The successful reconstruction of hip, knee, and ankle kinematic waveforms with high R^2^ values (0.981, 0.952, and 0.900, respectively) and reasonably low RMSE values (2.60°, 5.06°, and 6.01° respectively) indicates the robustness of the regression models. In addition, the RMSE values quantified the errors observed in the angular values of the reconstructed waveforms when compared to the measured angular values. The RMSE values for the hip and knee were below the critical 5° reported in literature, highlighting the fact that our model could be used for further predictions and analysis. This threshold value, >5° is considered significant and may have impact on the interpretation of gait analysis (Moissenet et al., 2019). The reconstructed ankle waveform deviated about 1°, resulting in an effect greater than the acceptable threshold. We hypothesized that the errors could be due to soft tissue artefacts observed in literature (Lavaill et al., 2022) or displacement of the ankle marker during the motion capture. This however require further investigation which could help improve our model in future research. We also proposed that the 1° deviation observed could be accounted for when using our models for further predictions.

The technique employed to reconstruct kinematic waveforms was based on fitting splines between different key events, which were estimated with linear, stepwise and robust regression model. Our model was able to predict and reconstruct the expected waveforms within one standard deviation of the mean reported in our dataset. The combined effect of the predictors were statistically significant after the stepwise regression analysis with WS as the predictor having the most effect on the *K*_*i*_ values (about 30%), similar to other works (Fukuchi et al., 2019). The significance of gender, BMI, and age were 17.20%, 9.68%, and 4.30% respectively. Also, interaction terms, representing the combined effect of pairs of predictors were significant 36 times (ie. 38.71%), regarding kinematics after performing stepwise regression. More importantly, the regressors allowed us to assess the relative contributions of the predictors to mismatches that commonly appear between a patient and normative database (Sabbagh et al., 2020). Our analysis revealed that BMI, age, gender, and walking speed significantly impact the kinematic waveforms. This aligns with previous research indicating that these variables are critical determinants of gait patterns (Abualait & Ahsan, 2022). For instance, walking speed has been shown to affect joint kinematics significantly, with faster speeds typically resulting in greater joint angles and velocities. Similarly, BMI influences the mechanical load on the joints, altering gait dynamics, and age-related changes in muscle strength and coordination further modify gait characteristics (Dewolf et al., 2021). Considering the narrow age range in our database (ie. 17 to 25 years), it is understandable that age was the less significant predictor for the model.

The use of a shape preserving spline algorithm for spline fitting and the application of leave-one-out cross-validation strengthened the robustness of our models. The choice of key points for waveform reconstruction, grounded in their biomechanical, p-values < 0.05 and clinical significance, ensured that the reconstructed waveforms were not only accurate but also meaningful from a clinical perspective (Hulleck et al., 2022).The development of these predictive models has substantial implications for clinical gait analysis in regions with similar demographic characteristics as those obtained in this study. The ability to generate accurate normative kinematic data specific to the Ghanaian population can enhance the precision of gait assessments, which could lead to better planning for region-specific CGA

While our study offers significant advancements, it is not without limitations. The sample size (*n* = 30), though adequate for initial modeling, should be expanded in future studies to enhance the generalizability of our findings. Additionally, while the current study focused on young adults, future research should consider other age groups to develop a comprehensive normative database across the lifespan. The integration of non-linear regression models may also enhance the predictive power and flexibility of our models, accommodating the complex interactions between multiple predictors and gait outcomes.

## 5 Conclusion

We successfully developed predictive models for the sagittal kinematics in young Ghanaian adults and demonstrated the significant impact of BMI, age, gender, and walking speed on gait patterns. These models represent a step toward personalized clinical gait analysis. Future research should aim to validate these models in larger and more diverse population, ensuring their applicability across different demographics and gait-related disorders. This would help clinicians to quantitatively make analysis for better diagnosis and treatment of gait disorders.

## Data Availability

All data produced in the present study are available upon reasonable request to the corresponding author.

https://github.com/PhilipKone/Predictive-Gait-Analysis-

## Abbreviations

CGA: Clinical gait analysis
LM: Linear Model
SLM: Stepwise Linear Model
RLM: Robust Linear Model
C1: self-selected normal walking speed
C2: slow walking speed
C3: fast walking speed
BMI: Body Mass Index
PCHIP: Piecewise Cubic Hermite Interpolating Polynomial
R^2^: coefficient of determination
RMSE: Root Mean Squared Error
WS: dimensionless walking speed
LED: Light Emitting Diode

## Acknowledgments

First off, we would like to thank the almighty God for a successful project. We would like to acknowledge and express our heartfelt gratitude and appreciation to the University of Ghana’s Department of Biomedical Engineering for providing as with the resources for data collection. We also express gratitude to Paul Normeshie and Mawuli Ahimbleame for their unwavering support. Finally, we would like to acknowledge all participants that volunteered to have their gait data for this research.

## References

Abdul Aziz Hulleck, Dhanya Menoth Mohan,Abdallah, N., Marwan El Rich, & Khalaf, K. (2022). Present and future ofgait assessment in clinical practice: Towards the application of novel trendsand technologies. Frontiers in Medical Technology, 4. 10.3389/fmedt.2022.901331

Abualait, T., & Ahsan, M. (2022). Comparisonof gender, age, and body mass index for spatiotemporal parametersof bilateral gait pattern. F1000Research, 10, 266. 10.12688/f1000research.51700.2

Al-kharaz, A. A., & Chong, A. K. (2022). Gender differences in ankle kinematics of adults during gait. Journalof Sex- and Gender-Specific Medicine, 8(3), 147–153. https://www.gendermedjournal.it/archivio/3927/articoli/39109/

Bas Huijben, Kimberley, Dieën, van, & Mirjam Pijnappels. (2018). The effect of walking speed on quality of gait inolder adults. Gait & Posture, 65, 112–116. 10.1016/j.gaitpost.2018.07.004

Bennett, H. J., Fleenor, K., & Weinhandl, J. T. (2018). A normative database of hip and knee jointbiomechanics during dynamic tasks using anatomical regression predictionmethods. Journal of Biomechanics, 81, 122–131. 10.1016/j.jbiomech.2018.10.003

Dewolf, A. H., Sylos-Labini, F., Cappellini, G.,Ivanenko, Y., & Lacquaniti, F. (2021). Age-related changes in theneuromuscular control of forward and backward locomotion. PLOS ONE, 16(2), e0246372. 10.1371/journal.pone.0246372

Eni Halilaj, Rajagopal, A., Madalina Fiterau, Hicks, J. L., Hastie, T., & Delp, S. L. (2018). Machine learning in human movement biomechanics: Bestpractices, common pitfalls, and new opportunities. Journal ofBiomechanics, 81, 1–11. 10.1016/j.jbiomech.2018.09.009

Florent Moissenet, Leboeuf, F., & Armand, S. (2019). Lower limb sagittal gait kinematics can be predicted basedon walking speed, gender, age and BMI. Scientific Reports, 9(1). 10.1038/s41598-019-45397-4

Fritsch, F. N., & Carlson, R. E. (1980). Monotonepiecewise cubic interpolation. SIAM Journal on Numerical Analysis, 17, 238–246.

Fukuchi, C. A., Fukuchi, R. K., & Duarte, M. (2019). Effects of walking speed on gait biomechanics in healthyparticipants: a systematic review and meta-analysis. Systematic Reviews, 8(1). 10.1186/s13643-019-1063-z

Galea, O., Bristow, H. D., Chisholm, S.M., Mersch, M. E., Nullmeyer, J., Reid, C. R., & Treleaven, J. (2019). Single and dual tandem gait assessment post-concussion: What performance timeis clinically relevant across adult ages and what can influence results? MusculoskeletalScience and Practice, 42, 166–172. 10.1016/j.msksp.2019.04.006

Gross, R., Robertson, J., Leboeuf, F.,Hamel, O., Brochard, S., & B. Perrouin-Verbe. (2017). Neurotomy of therectus femoris nerve: Short-term effectiveness for spastic stiff kneegait. Gait & Posture, 52, 251–257. 10.1016/j.gaitpost.2016.11.032

Hagoort, I., Vuillerme, N., TiborHortobágyi, & Claudine. (2022). Outcome-dependent effects of walking speedand age on quantitative and qualitative gait measures. Gait & Posture, 93, 39–46. 10.1016/j.gaitpost.2022.01.001

Hof, A. L. (1996). Scaling gait data to body size. Gait & Posture, 4(3), 222–223. 10.1016/0966-6362(95)01057-2

Hulleck, A. A., Menoth Mohan, D., Abdallah, N., ElRich, M., & Khalaf, K. (2022). Present and future of gait assessment inclinical practice: Towards the application of novel trends and technologies. Frontiersin Medical Technology, 4. 10.3389/fmedt.2022.901331

Kahaner, D., Moler, C., & Nash, S. (1988). Numericalmethods and software. Upper Saddle River, NJ: Prentice Hall.

World Medical Association. (1974). WMA Declarationof Helsinki: ethical principles for medical research involving human subjects.353 (1), 1418–1419. http://www.wma.net/en/30publications/10policies/b3/index.html

Karimi, G., & Jahani, O. (2012). Genetic AlgorithmApplication in Swing Phase Optimization of AK Prosthesis with Passive Dynamicsand Biomechanics Considerations. Genetic Algorithms in Applications. 10.5772/38211

Kaya, E. (2014). Spline Interpolation Techniques. Journal of Technical Science andTechnologies, 47–52. 10.31578/jtst.v2i1.56

Lavaill, M., Martelli, S., Kerr, G. K., & Pivonka, P. (2022). Statistical Quantification of the Effects of Marker Misplacement andSoft-Tissue Artifact on Shoulder Kinematics and Kinetics. Life, 12(6), 819. 10.3390/life12060819

Lu, T.-W., & Chang, C.-F. (2012). Biomechanics ofhuman movement and its clinical applications. The Kaohsiung Journal ofMedical Sciences, 28(2), S13–S25. 10.1016/j.kjms.2011.08.004

Mikos, V., Yen, S.-C., Tay, A., Heng, C.-H., Chung, C.L. H., Liew, S. H. X., Tan, D. M. L., & Au, W. L. (2018). Regressionanalysis of gait parameters and mobility measures in a healthy cohort forsubject-specific normative values. PloS One, 13(6), e0199215. 10.1371/journal.pone.0199215

M.J. Booij, Richards, R., Jaap Harlaar, & van. (2020). Effect ofwalking with a modified gait on activation patterns of the knee spanningmuscles in people with medial knee osteoarthritis. The Knee, 27(1), 198–206. 10.1016/j.knee.2019.10.006

Moissenet, F., Leboeuf, F., & Armand, S. (2019). Lower limb sagittal gait kinematics can be predicted based on walking speed,gender, age and BMI. Scientific Reports, 9(1). 10.1038/s41598-019-45397-4

Obesity and Osteoarthritis: ABiomechanical and Global Health Evaluation of Total Knee Arthroplasty Patientsand Healthy Controls - ProQuest. (2021). Proquest.com. https://www.proquest.com/openview/72136df01e18439c7664742b7ddf6a58/1?pq-

Pietraszewski, B., Winiarski, S., & Jaroszczuk, S. (2012). Three-dimensional human gait pattern - Reference data for normal men. Actaof Bioengineering and Biomechanics, 14(3), 9–16. 10.5277/abb120302

Porta, M., Pau, M., Leban, B., MichelaDeidda Sorrentino, M., Arippa, F., & Marongiu, G. (2021). Lower Limb Kinematicsin Individuals with Hip Osteoarthritis during Gait: A Focus on AdaptativeStrategies and Interlimb Symmetry. Bioengineering, 8(4), 47–47. 10.3390/bioengineering8040047

Rowe, E., Beauchamp, M. K., & Wilson, J. (2021). Age and sex differences in normative gait patterns. Gait & Posture, 88, 109–115. 10.1016/j.gaitpost.2021.05.014

Sabbagh, D., Ablin, P., Gaël Varoquaux,Alexandre Gramfort, & Engemann, D. A. (2020). Predictive regressionmodeling with MEG/EEG: from source power to signals and cognitive states. NeuroImage, 222, 116893–116893. 10.1016/j.neuroimage.2020.116893

Schreiber, C., & Florent Moissenet. (2019). A multimodal dataset of human gait at different walking speeds established oninjury-free adult participants. Scientific Data, 6(1). 10.1038/s41597-019-0124-4

Strongman, C., & Morrison, A. P. (2020). A scoping review ofnon-linear analysis approaches measuring variability in gait due to lower bodyinjury or dysfunction. Human Movement Science, 69, 102562–102562. 10.1016/j.humov.2019.102562

Sun, D., Fekete, G., Mei, Q., & Gu, Y. (2018). The effect of walking speed on the foot inter-segment kinematics,ground reaction forces and lower limb joint moments. PeerJ, 6, e5517–e5517. 10.7717/peerj.5517

Vervoort, D. (2020). Adaptability of gaitand balance across the adult lifespan. University of Groningen ResearchDatabase (University of Groningen/ Centre for Information Technology). 10.33612/diss.144620201

Xu, L., Xu, W., Golyanik, V., Habermann, M., Fang, L., & Theobalt, C. (2020). EventCap: Monocular 3D Capture of High-Speed HumanMotions Using an Event Camera. Thecvf.com, 4968–4978.https://openaccess.thecvf.com/content_CVPR_2020/html/Xu_EventCap_Monocular_3D_Capture_of_High-Speed_Human_Motions_Using_an_CVPR_2020_paper.html

